# The novel CFAbd-Score.kid^©^ reveals a significant decline of abdominal symptoms in children with Cystic fibrosis aged 6 through 11 years on Elexacaftor/Tezacaftor/Ivacaftor – first results

**DOI:** 10.1101/2024.10.29.24316355

**Authors:** Jochen G. Mainz, Pauline Sadrieh, Lilith Bechinger, Franziska Duckstein, Anton Barucha, Louise Polte, Lutz Naehrlich, Olaf Eickmeier, Suzanne van Dullemen, Ute Graepler-Mainka, Carlos Zagoya

## Abstract

**Background:** Recently, elexacaftor/tezacaftor/ivacaftor (ETI), the turning point in the course of Cystic fibrosis (CF), was also approved for children with CF (cwCF) aged 6-11years carrying at least one F508del-mutation. Modulating the causative deficiency in the CF transmembrane conductance regulator channel was found to substantially improve the crucially affected respiratory and digestive CF-manifestations. In this regard, for people with CF aged ≥12years, we previously found that ETI decreases significantly abdominal symptoms (AS) using the CFAbd-Score^©^.

**Aims:** Assessing changes in AS after ETI initiation in cwCF aged 6-11years with the novel pediatric Patient-Reported Outcome Measure CFAbd-Score.kid^©^.

**Methods:** The CFAbd-Score.kid^©^, specially developed for cwCF aged <12years, implements pictograms, easy language and children-oriented response strategies, comprising 29 CF-specific gastrointestinal items from five domains. Its scoring algorithm developed following FDA guidelines weights items and domains differently, reaching a maximum of 100 points. CwCF completing at least one questionnaire before ETI initiation and another one during ETI therapy were included.

**Results:** In four German CF centers, a total of n=52 cwCF (mean age 8.3±2.2years) were included, completing a total of n=293 questionnaires. During ETI therapy, significant decreases were observed for mean total CFAbd-Score.kid^©^ (−31%/p<0.0001) as well as for mean sub-scores of “pain” (−26%/p<0.01), “QoL impairment” (−48%/p<0.01), “disorders of bowel movement” (−32%/p≤0.0001) and “disorders of appetite” (−42%/p<0.05).

**Conclusion:** Among cwCF aged 6-11years, AS captured with the novel CFAbd-Score.kid^©^ significantly decreased during the novel ETI treatment. Simultaneously, CFAbd-Score.kid^©^ proved to be sensitive to ETI-induced changes in AS. Further validation steps and international implementation are currently in progress.

## 1 Introduction

Highly effective modulator therapy (HEMT) elexacaftor/tezacaftor/ivacaftor (ETI), which proved to be a turning point in the course of Cystic fibrosis (CF) in people with CF (pwCF) aged ≥12 years, was also recently approved for children with CF (cwCF) aged 6 through 11 years (1). Targeting the causative molecular defect in the CF transmembrane conductance regulator (CFTR) channel (2), HEMTs have substantially changed the course of the multi-organ disease, which principally affects the respiratory and digestive systems.

Mutations in the *CFTR* gene lead to dysfunctional CFTR channels in the apical membranes of the exocrine glands, which impairs the outflow of chloride ions, resulting in osmotic imbalances and secretions with increased viscosity (3, 4). ETI modulates the CFTR channel defect by increasing the availability and functionality of the CFTR protein on the cell surface as well as the opening probability of the CFTR channel, resulting in improved chloride transport and, thus, reducing the viscosity in the secretions from the exocrine glands (5). Accordingly, HEMT has proved to markedly decrease sweat chloride concentrations in adults and cwCF (6, 7).

Since pulmonary involvement remains to be the main cause of premature death in pwCF (3), studies have mainly focused on the pulmonary impact of ETI therapies (8). Accordingly, primary endpoints in HEMT trials in adult and cwCF were changes in pulmonary function as measured by FEV_1_%pred and in the lung clearance index, which revealed marked and significant improvements (6, 7).

The burden of abdominal symptoms (AS) deriving from a CF specific pattern of multi-organ GI-involvement in pwCF (9) has been found to have a substantial impact on the quality of life (QoL) in pwCF (2). Two studies focusing on the effects of ETI therapy on AS using the CFAbd-Score^©^ in pwCF ≥12 years recently showed a clinically meaningful and statistically significant decrease in the burden of AS (8, 10). The question proposed was whether this holds also for 6– 11-year-old cwCF.

Since there were no specific patient-reported outcome measures (PROMs) for cwCF, based on the CFAbd-Score^©^, we developed the CFAbd-Score.kid^©^ questionnaire, which is currently undergoing a comprehensive validation process(11). Following FDA and EMA guidelines as well as COSMIN and ISPOR recommendation (12–15), we consulted focus groups with cwCF, their proxies, as well as CF specialists from different fields during the development of the pediatric PROM. The resulting 29 items included in the CFAbd-Score.kid^©^ refer to the specific pattern of AS reported by cwCF, including flatulence, impaired appetite and abdominal pain, as well as the impact of AS on the patients’ psychosocial wellbeing. Due to the implemented pictograms and child-friendly language, cwCF aged 6-11 years were able to complete the questionnaire independently or with support of their proxies. Originally developed for ‘augmentative and alternative communicatioߣ, the implemented pictograms were provided by the platform Metacom^©^ (16), which were selected and adapted for inclusion in the CFAbd-Score.kid^©^. They support the children’s comprehension of the corresponding sentences and facilitate recognition of questions in terms of repeated/iterative questionnaire administrations (17).

The aim of the study was to assess changes in AS of cwCF aged 6-11 years during the implementation of a novel treatment with ETI, which, previous to this study, had been approved in Germany for this age group. A further aim was to validate the new pediatric PROM in regard to “sensitivity to change” induced by the new therapy.

## 2 Methods

### 2.1 Ethical statement

This study has been conducted in strict accordance with the ethical guidelines of the Declaration of Helsinki and was approved by the Brandenburg Medical School (MHB) ethics committee (registration number E-01-20200519). Furthermore, it has been registered in ClinicalTrials.gov identifier: NCT03052283, Development and Validation of a Disease Specific PROM to Assess Abdominal Involvement in pwCF (CFAbd-Score^©^ / CFAbd-Score.kid^©^).

### 2.2 Participants

CwCF aged between 6-11 years were recruited at four CF centers in Germany (Brandenburg an der Havel, Tübingen, Giessen and Frankfurt am Main). Inclusion criteria were confirmed evidence of CF by two positive sweat tests and/or CF genotyping, as well as the presence of at least one F508del mutation, following ETI-approval conditions. Exclusion criteria were non-eligibility for ETI therapy, as well as the lack of a minimum of two completed questionnaires, with at least one required to be completed prior to ETI initiation and another one during ETI therapy. Pulmonary function (FEV_1_%pred), airway colonization with specific pathogens and non-GI comorbidities were reported but not an exclusion criteria. Patients and their parents/legal guardians provided written informed consent.

### 2.3 Abdominal symptom assessment and the CFAbd-Score.kid^©^

The CFAbd-Score.kid^©^ applied to capture and quantify AS before and during ETI therapy, has been developed based on the validated CFAbd-Score^©^ (18, 19). It was designed to enable children below the age of 12 years to record their GI symptoms mostly independently or with little support from their parents and/or guardians, using a combination of easy/child-friendly language and pictograms selected and adapted from the platform Metacom^©^ (16, 17) to facilitate understanding and the ability to respond adequately (Fig.1). Altogether, the CFAbd-Score.kid^©^ consists of 29 CF-specific items grouped into 5 domains: pain (5 items), gastroesophageal reflux disease (GERD/upper GI) (4 items), quality of life impairment (QoL) (9 items), disorders of bowel movement (DBM/lower GI) (7 items) and disorders of appetite (DA) (4 items). Analogous to the version intended for pwCF >12, the CFAbd-Score.kid^©^ includes a modified Bristol Stool Shape Scale as well as a color stool chart. Being a retrospective tool, questions are answered recalling the past two weeks. Additionally, basic demographic and medical information about the patients were collected in this study. In order to collect additional general information such as age, sex, enzyme intake, the information sheet was accordingly adapted for this age group.

**Figure 1.**
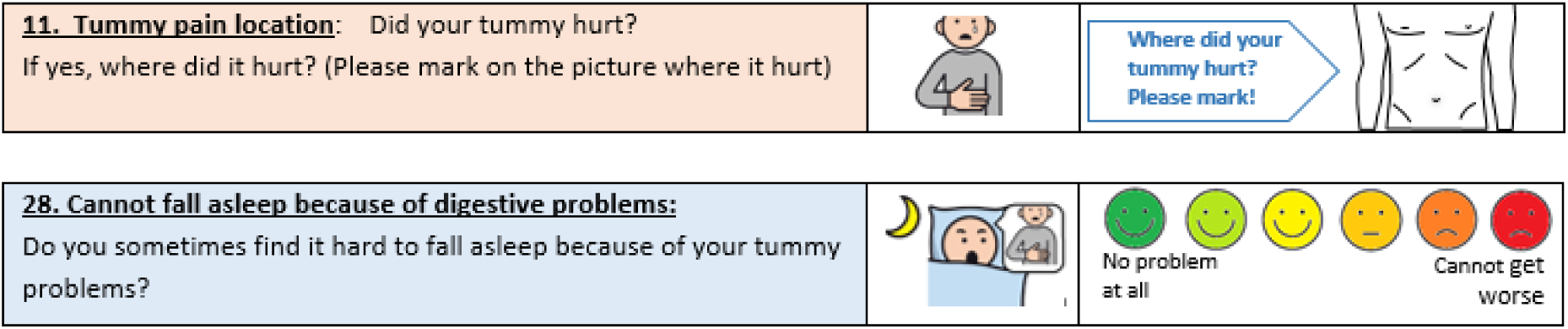
Exemplary pictograms and questions implemented in the CFAbd-Score.kid^©^ translated into English by Franziska Duckstein (MA, professional translator).

### 2.4 Statistical analysis

Changes in the total CFAbd-Score.kid^©^ and its five domains were assessed including all questionnaires with at least 50% of answers completed before and after ETI initiation. Since some participants provided multiple questionnaires corresponding to different time points previous to and during ETI therapy, linear mixed-effects models (LMEMs) with random slopes were used to account for within-subject correlation. The intra-class correlation coefficient (ICC) computed for the both the intercept and slope indicated that approximately 86% of the variance was accounted for by allowing the model’s intercept and slope to vary across individuals. These models included the variables sex (f/m), use of previous CFTR modulators (yes/no) and genotype (homozygous/heterozygous for F508del) as fixed effects. Normality assumptions on LMEM’s residuals were verified by Q-Q plots and visual inspection of residuals distributions. Data from domains “GERD/upper GI”, “disorders of appetite” and “quality of life impairment” was log-transformed to satisfy normality assumptions on LMEM’s residuals. Results from CFAbd-Score.kid^©^ are reported as estimated marginal means and standard error of mean in the original scale, as computed with the R *lsmeans* package. Exploratory analyses were conducted to assess ETI-induced changes in 28 of the 29 items included in the CFAbd-Score.kid^©^.

Changes in the percentages of cwCF reporting GI symptoms with frequency ≥2 days per 2 weeks or with severity of at least 2 points in a 6-point Likert scale were calculated using two questionnaires per patient, i.e. the most recent questionnaire previous to ETI therapy and the last questionnaire completed during ETI therapy. Furthermore, we defined stool frequency as suspicious when patients reported <4 stools/14 days or >21 stools/14 days, despite not necessarily considered to be pathologic. Stool consistency was considered irregular when patients reported having hard or loose consistency stools, as assessed with a modified Bristol Stool Shape Scale included in the CFAbd-Score.kid^©^.

In order to identify age-related differences in ETI-induced effects on GI symptoms, these changes were compared with the changes in a subgroup of German pwCF aged ≥12 years reported with the CFAbd-Score^©^ in a study published in 2023 (20). BMI-for-age z-scores were calculated using the LMS method using growth reference data from the World Hearth Organization (21). Changes in BMI-for-age z-scores and pulmonary function FEV_1_%pred were assessed using student’s t-tests for paired samples and two data points per patient (previous to and during ETI therapy) for each of these parameters. All statistical analyses were performed using R version 3.6.3.

## 3 Results

### 3.1 Demographics and clinical characteristics

A total of 293 CFAbd-Score.kid^©^ questionnaires collected from n=52 cwCF attended in the four participating CF centers (Brandenburg an der Havel n=8, Frankfurt n=13, Tübingen n=23 and Giessen n=8) were evaluated in the study. At ETI therapy start, all patients included in the study were aged between 6-11 years, according to the drug approval criteria. However, at the time of the baseline assessment for inclusion before starting ETI, mean age was 8.3±2.2 years (min: 4.7, max: 11.7 years). Altogether, n=137 CFAbd-Score.kid^©^ questionnaires were collected previous to ETI initiation and n=156 during ETI therapy, which, throughout the entire observation time frame, resulted in a median of 6 questionnaires (IQR: 4, 7) per patient. Table 1 provides details about demographics and clinical characteristics of the included cwCF.

**Table 1.** Baseline demographics and clinical characteristics of the included cwCF aged 6-11 years.

|  |  | **Total number**<br>**of cwCF (n, %)** |
| --- | --- | --- |
| **Sex** | female | 36 (69) |
|  | male | 16 (31) |
| **Genotype** | F508del homozygous | 30 (58) |
|  | F508del heterozygous | 22 (42) |
| **Previous**<br>**CFTR modulator**<br>**therapy** | yes | 26 (50) |
|  | LUMA/IVA | 25 (48) |
|  | IVA | 1 ( 2) |
|  | no | 26 (50) |
| **Exocrine**<br>**pancreatic insufficiency** | yes | 50 (96) |
|  | no | 2 ( 4) |
| **FEV<sub>1</sub> at baseline** | $93.5 \pm 19.4$ %pred | |
| **BMI-for-age z-score** | $-0.46 \pm 1.0$ | |

According to the inclusion criteria, each of the 52 cwCF included completed at least two questionnaires, one before ETI initiation (median, IQR: −16 [−100, −1] days) and another one during ETI therapy (median, IQR: 160 [52, 283] days). After commencing ETI therapy, 47/52 (90%) filled out at least one questionnaire within the first 6 months (24 weeks), and the remaining 5/52 (10%) by month 21(84 weeks) on the new therapy. Only 5 questionnaires were completed by 5 children within the first two weeks (14 days) of therapy with ETI. For single-item analysis, changes in symptom frequencies were assessed with 136 CFAbd-Score^©^ questionnaires from 68 German pwCF aged ≥12 years (median age [IQR]: 19 [14, 24] years; min, max: 12, 55 years), i.e. two per patient: one before and one after the start of ETI (8).

### 3.2 Changes in the Total CFAbd-Score.kid^©^ and its five domains

During ETI therapy, significant decreases were observed for the mean total CFAbd-Score.kid^©^ (−38%, p<0.0001) as well as for mean sub-score domains “Pain” (−37%, p<0.0001), “QoL impairment” (−45%, p<0.001), “DBM/Lower GI” (−31%, p<0.0001) and “DA” (−57%, p<0.001) (Fig. 2 and Tab. 2).

**Figure 2.**
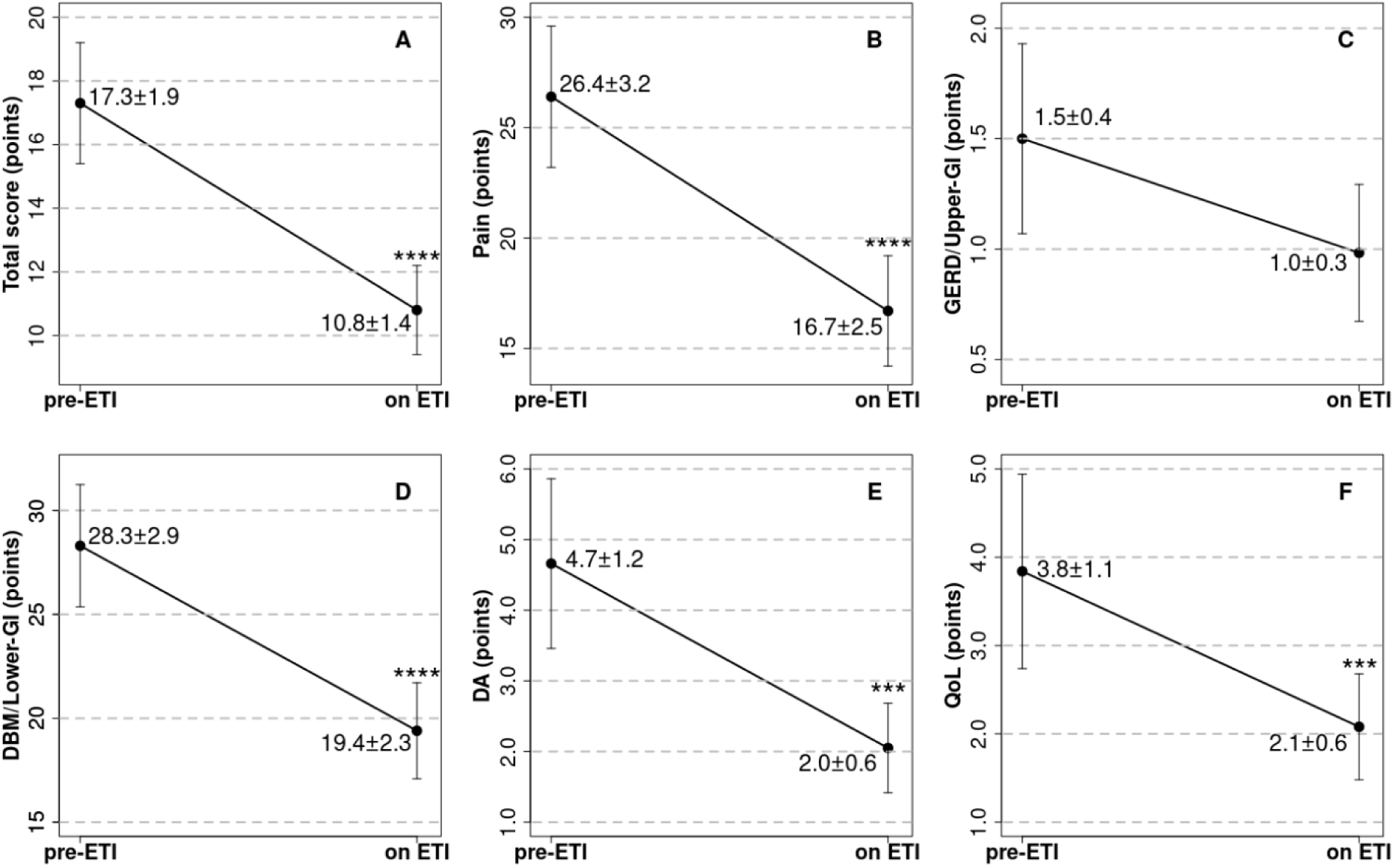
Changes in means (±SEM) of the total CFAbd-Score.kid*^©^* (A) and its five domains: (B) pain, (C) GERD/upper GI, (D) disorders of bowel movement (DBM/lower GI), (E) disorders of appetite (DA) and (F) quality of life (QoL), after implementation of a new ETI therapy. Statistical significance is represented with the symbols *, **, ***, and ****, for p<0.05, p<0.01, p<0.001, and p<0.0001, respectively.

**Table 2.**
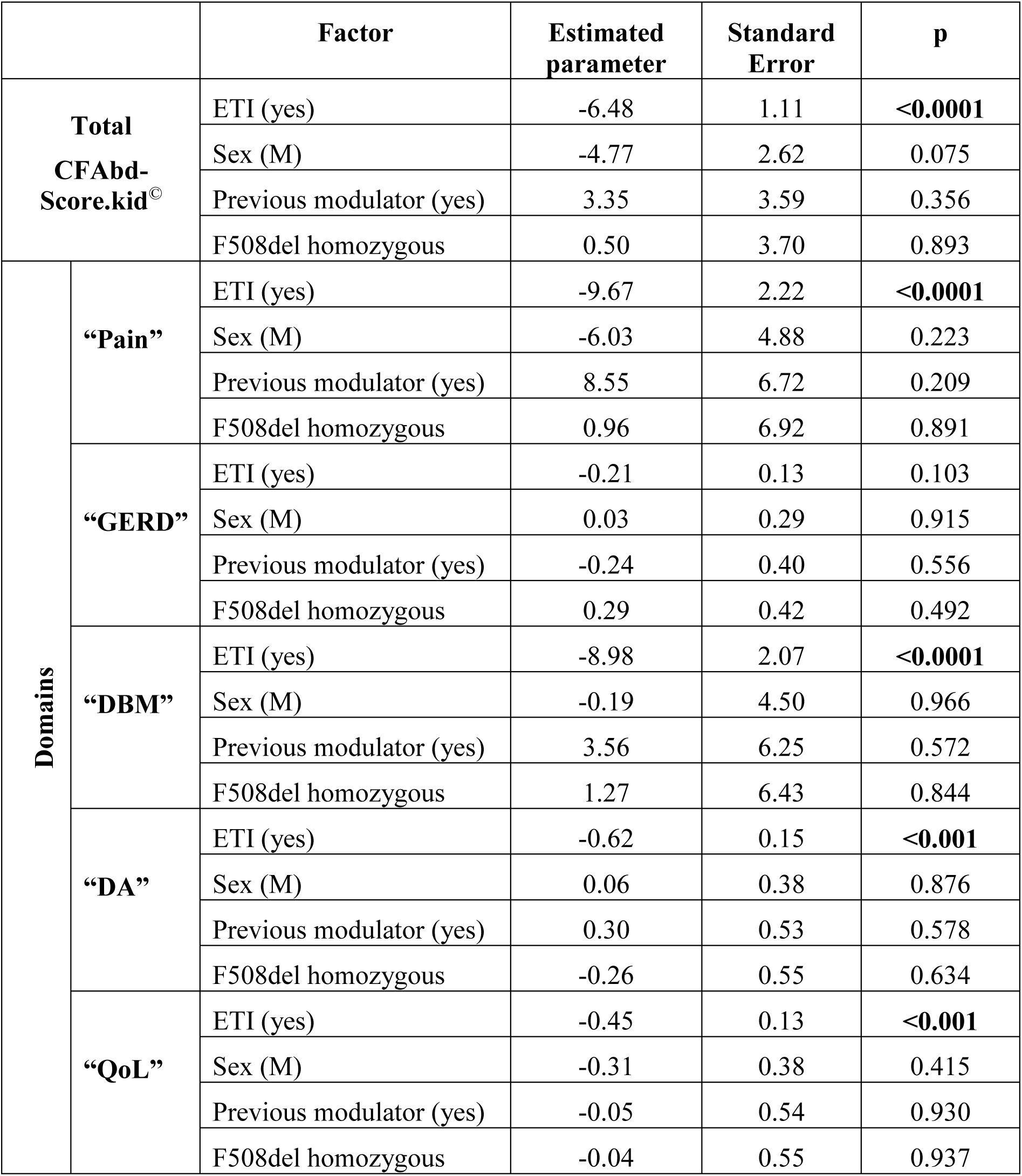
Parameters resulting from the linear mixed-effects models for the total and its five domains: “pain”, “GERD”, “disorders of bowel movement (DBM)”, “disorders of appetite (DA)” and “quality of life impairment (QoL)”.

Treatment with ETI prompted a significant reduction in GI symptoms in total CFAbd-Score.kid^©^ score and its domains pain, disorders of bowel movement (DBM/lower GI), disorders of appetite (DA) and GI-related quality of life (QoL). Declines in domain GERD/upper GI, which, at baseline, accounted for the lowest burden of symptoms (only 1.5 points) were not significant. Neither were there significant differences in the subgroups like sex, genotypes or previous CFTR modulator therapy (Tab. 2). However, the total CFAbd-Score.kid^©^ and the domains “Pain” and “DBM/lower GI” from questionnaires completed after 24 weeks on ETI therapy (n=79, median time: 283 [210, 354] days) were significantly lower, compared to questionnaires (n=77, median time: 50 [(20)27, 102] days) completed within the first 24 weeks (total: 12.4±1.6 points → 9.8±1.5 points; Pain: 20.3±3.1 points → 13.3±2.8 points; DBM/lower GI: 21.5±2.8 points → 15.7±2.1 points; all p<0.05).

In Figure 3, the distribution of the most recent questionnaire completed prior to ETI initiation is shown in regard to its quartiles. The first, second and third quartiles of the respective distribution of scores resulted in Q_1_=9.1 points, Q_2_=15.3 points and Q_3_=23.4 points (Fig.3). The maximum score was 52.7/100 points.

**Figure 3.**
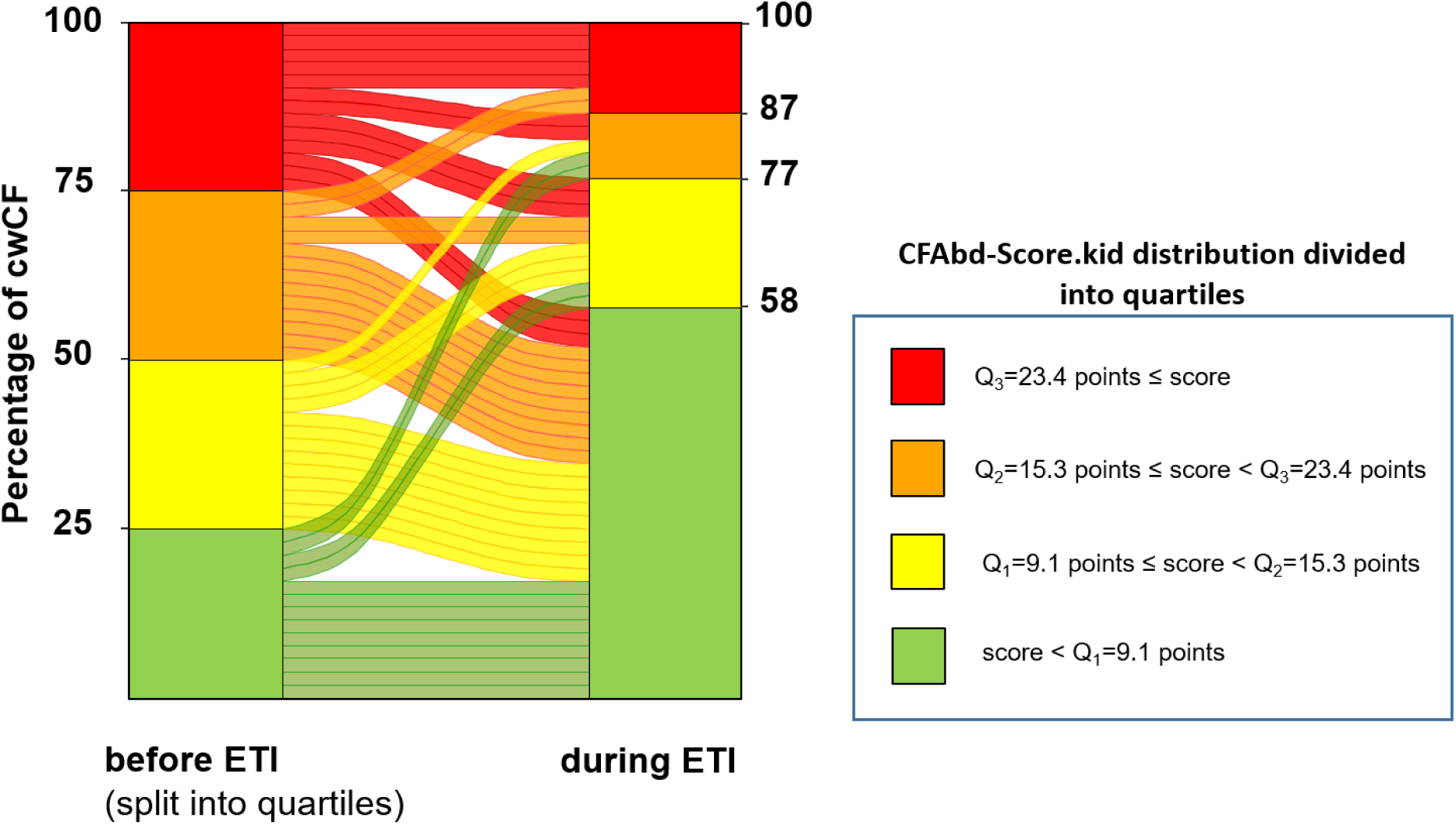
Dynamics of the distribution of CFAbd-Score.kid^©^ scores (n=52) in regard to their severity displayed as quartiles of the pre-ETI distribution. Q1=9.1 points, Q2=15.3 points, and Q3=23.4 points represent the first, second and third quartiles of the pre-ETI distribution, respectively. Baseline CFAbd-Score.kid^©^ has been distributed into quartiles.

The dynamics of AS severity for each patient (Fig.3) revealed that during ETI therapy, 58% (30 patients) of the patients’ scores lay below the baseline quartile of Q_1_=9.1 points. Another 19% (10 patients) lay between Q_1_=9.1 points and Q_2_=15.3 points, and 10% (5 patients) between Q_2_=15.3 and Q_3_=23.4 points. Unlike the pre-ETI distribution of scores, only 13% (7 patients) scored more than Q_3_=23.4 points in the CFAbd-Score.kid^©^, i.e. a 46% reduction in the percentage of patients reporting GI symptoms above Q_3_, compared to the baseline distribution. At the same time, the maximum score of the distribution (reached by one of the included children) declined from 52.7 to 41.5/100 points.

Interestingly, the burden of AS increased in a smaller number of patients. Namely, four of the 13 patients initially scoring below Q_1_ revealed an increase in their burden of GI symptoms after ETI initiation: 2 above Q_2_, and 2 others above Q_3_, according to the score distribution before ETI initiation (Fig.3).

### 3.3 Frequency of symptoms assessed with individual items of the CFAbd-Score.kid^©^ before and after ETI initiation

The percentage of items responses in the CFAbd-Score.kid^©^ showing frequent and/or severe symptom burden, before ETI initiation, in the included children (see definition in the Methods section) declined during the new therapy with ETI. Only the proportion of responses for the items “vomiting” and “constipation” did not reveal changes. For the item reporting the symptom of “reflux”, initially reported only scarcely, the proportion of responses resulted to increase by 4% (Fig.4 and 5).

**Figure 4.**
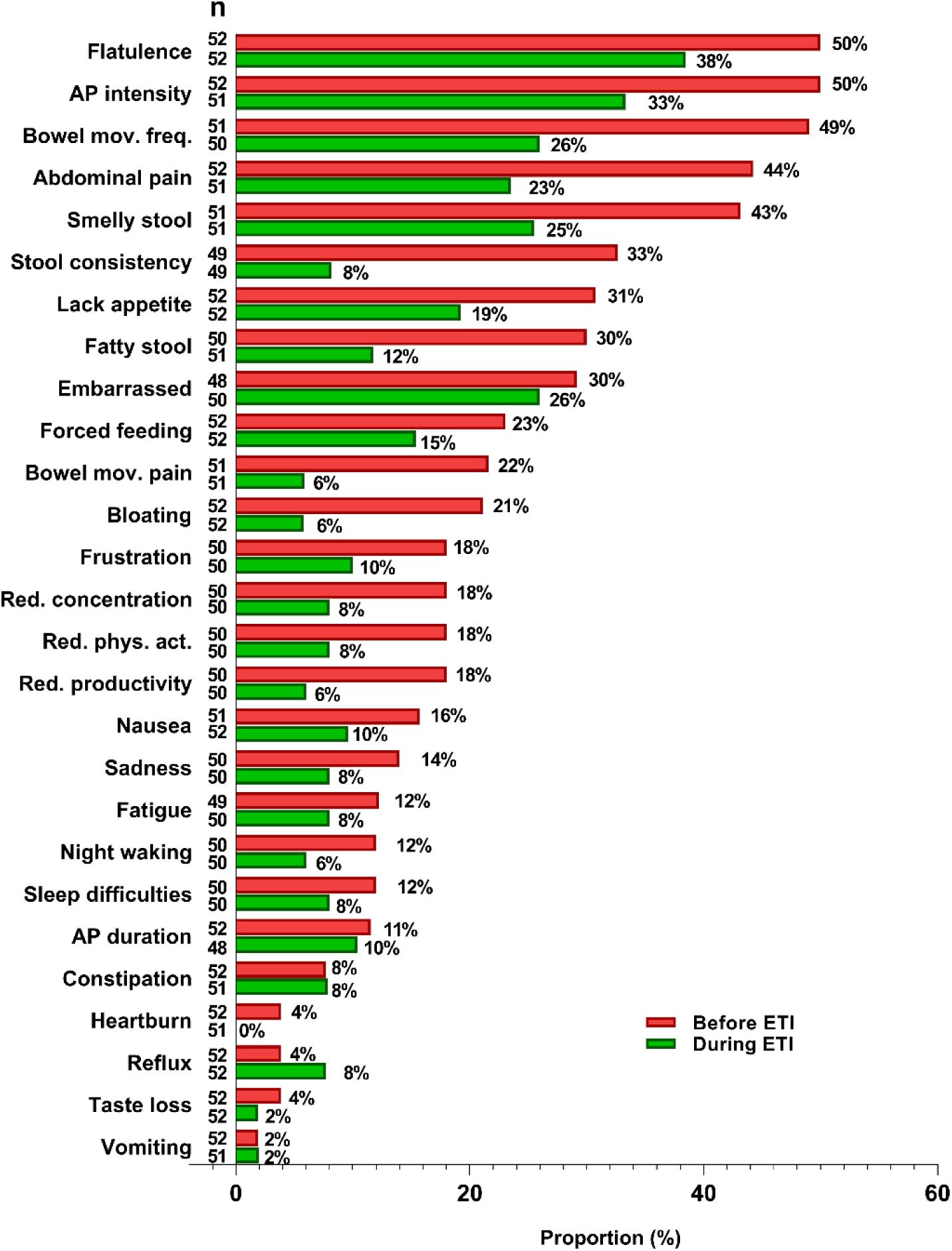
Percentages of cwCF reporting GI symptoms as recorded with 27 out of the 29 items of the CFAbd-Score.kid*^©^* before and during ETI therapy. Percentages represent the proportion of cwCF reporting the corresponding item with frequency ≥2-3 times or with severity of at least 2 points in a 6-point Likert scale, during the 2-week recall time frame. Numbers in the column on the right-hand side of each item name (n) represent the number of total answers for the respective item.

Before ETI initiation, “flatulence” was the symptom for which 50% of cwCF, the highest percentage, reported experiencing this symptom at least twice during the past 2 weeks. Analogously, “abdominal pain intensity” was reported by 50% of the cwCF before ETI initiation with severity ≥2 in a 6-point Likert scale. During ETI, the proportion of children reporting these symptoms decreased by more than 10% (Fig.4). Other symptoms reported by more than 25% of the cwCF prior to ETI initiation were suspicious “bowel movement frequency”, “abdominal pain”, “smelly stool”, irregular “stool consistency”, “lack of appetite”, “fatty stool” and “embarrassment about GI symptoms”. The number of cwCF experiencing these symptoms decreased by more than 15% during ETI, except for “lack appetite” and “embarrassed” (Fig.4 and Fig.5).

**Figure 5.**
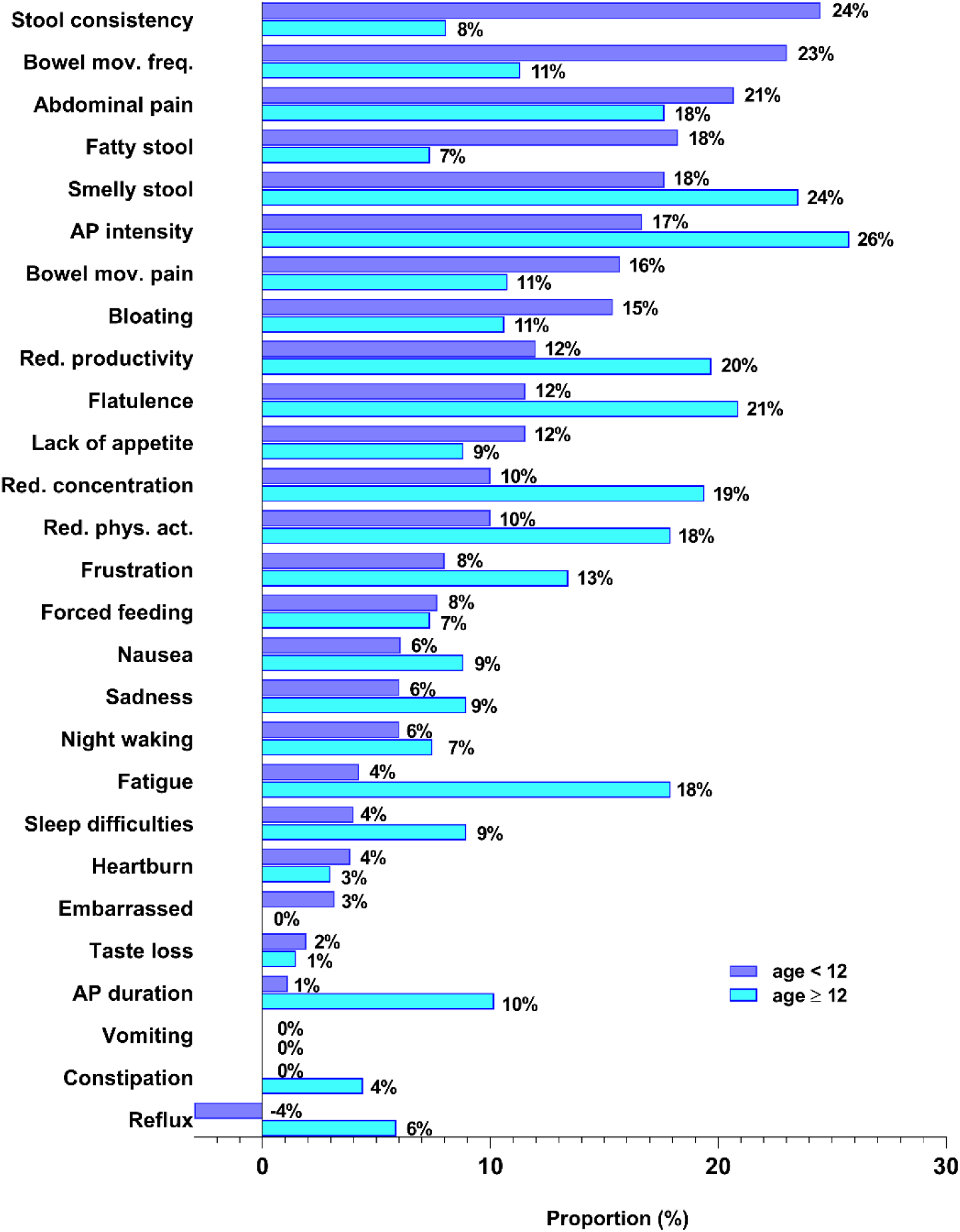
Comparison of ETI-induced changes in symptom frequency in cwCF aged 6 through 11 years, captured with the CFAbd-Score.kid*^©^*, with a previously published study cohort of pwCF aged ≥ 12 years, captured with the CFAbd-Score*^©^* (8).

Out of the 29 GI-related symptoms reported by cwCF with the CFAbd-Score.kid**^©^**, “stool consistency” was the symptom undergoing the greatest change during ETI, followed by “bowel movement frequency”. We defined ‘suspicious stool frequenc’ when patients reported <4 stools/14 days or ≥21 stools/14 days, despite not related to any definitions of constipation or diarrhea (Fig.3 and 4). Other symptoms for which the proportion of pwCF decreased by at least 10% were: “abdominal pain”, “fatty stool”, “smelly stool”, “abdominal pain intensity”, “pain during bowel movements”, “bloating”, “reduced productivity”, “flatulence”, “lack of appetite”, “reduced concentration” and “reduced physical activity”.

The percentage of pwCF reporting GI-related symptoms such as “fatigue”, “sleep difficulties”, “heartburn”, “embarrassed”, “taste loss”, “vomiting” and “constipation” did not surpass the 5% level, which also regarded “abdominal pain duration”. The percentage of cwCF reporting “reflux” increased by 4% after initiation of ETI (Fig.4).

### 3.4 Exploratory comparison of ETI-induced changes in GI symptom frequency in cwCF aged 6 through 11 (CFAbd-Score.kid^©^) and in pwCF aged ≥12 years (CFAbd-Score^©^)

Although the total scores in both cohorts declined with high significance, an exploratory comparison between the present cohort of children aged 6-11 years and pwCF aged ≥12 years, assessed with the CFAbd-Score^©^ in our previously published study (8), revealed some interestingly different effects of ETI in regard to the age group (Fig.5). Highest differences in its effects on abdominal symptoms were observed regarding the 4 items: “stool consistency”, “fatigue”, “bowel movement frequency” and “fatty stool”. Whereas the symptom “stool consistency” underwent the greatest change in the younger cohort, the change in the older cohort of pwCF did not surpass 8%. Furthermore, although there was a change of 23% in the proportion of younger pwCF reporting improvement in their “bowel movement frequency”, the correspondingly resulting change in the older cohort was of only 11% (Fig.4).

On the other hand, whereas the percentage of pwCF in the older cohort reporting “fatigue” improved by 18%, the respective percentage in the younger cohort using the CFAbd-Score.kid^©^ only improved by 4%. Analogously, the change in the percentage of pwCF reporting “reduced concentration” in the older cohort was 19%, whereas in the younger cohort it was 10%.

Differences of 5% to 10% between both age groups and questionnaires were found regarding “flatulence”, “abdominal pain intensity”, “abdominal pain duration”, “reduced physical activity”, “reduced productivity”, “smelly stool”, “frustration”, “sleep difficulties”, “bowel movement” and “bloating”. Finally, differences did not exceed 5% for “constipation”, “embarrassed”, “abdominal pain”, “sadness”, “nausea”, “lack of appetite”, “reflux”, “waking up at night”, “heartburn”, “taste loss”, “forced feeding” and “vomiting”.

### 3.5 Changes in pulmonary function and BMI-for-age z-scores

A significant improvement was observed in pulmonary function during ETI therapy, as measured by FEV_1_%pred, available in n=39 cwCF aged 6 through 11 years. FEV_1_%pred increased by 7.9%pred from a mean of 93.5 ± 19.4%pred to 101.5 ± 12.2%pred (p<0.001; median time (IQR) on ETI: 238, 98-337 days).

A significant improvement in the relative weight was observed in BMI-for-age z-scores, a pediatric age- and sex-adjusted measure, available in n=50 cwCF at baseline (median: −1day, IQR: [−7, 0]) and during therapy (median: 31days, IQR: [26, 93]). BMI-for-age z-scores improved by 49% from −0.46 ± 1.0 to −0.27 ± 0.94 (p=0.02).

## Discussion

Abdominal symptoms are a hallmark of CF, relevantly burdening and reducing patients’ quality of life (22–24). The increased life expectancy of pwCF is bringing the diagnosis and treatment of abdominal manifestations in pwCF into the clinical and scientific focus (23, 24). Accordingly, understanding “how can we relieve gastro-intestinal symptoms, such as stomach pain, bloating and nausea?” has been highlighted as one of the top ten priorities by the James Lind Alliance in 2018 and its updated survey from 2023 (25, 26). Furthermore, in the most recent survey, the newly identified priority “What are the effects of modulators on systems outside the lungs such as pancreatic function, liver disease, gastro-intestinal, bone density etc.?” ranked number 7. Nevertheless, most research regarding new therapies has been focused on the pulmonary manifestations of the multi-organ disease (19). To some extent, we attribute this to the previous lack of reliable PROMs focusing on the GI manifestations of CF (22, 24). Moreover, with the approval of ETI extended to cwCF aged 6 through 11 years, the question arises whether ETI may also decrease AS in cwCF, as we have previously shown for pwCF aged ≥12 years (8, 10, 20).

The present publication reveals, for the first time, analogous improvement in abdominal symptoms before and after ETI initiation in cwCF aged <12 years, primarily implementing the novel CFAbd-Score.kid^©^, whose validation is currently in progress (11). This novel CF-specific pediatric GI-PROM, has been developed based on the CFAbd-Score^©^ according to FDA and EMA guidelines as well as considering the COSMIN and ISPOR recommendation checklists (12–15). It enables pediatric patients below the age of 12 years to record their GI symptoms mostly independently, using a combination of child-friendly language, child-oriented response strategies and pictograms selected and adapted from the Metacom^©^ platform (Fig. 1). The total CFAbd-Score.kid^©^ is calculated using a proprietary scoring algorithm that weights each item and domain differently, reaching a maximum of 100 points corresponding to the highest burden of GI symptoms.

At present, only two projects examining the effects of ETI on AS have been published. The first and larger one being the North-American GALAXY/PROMISE study (22, 27, 28), using three gastrointestinal questionnaires and one “stool-specific questionnaire” as an online tool, developed and validated for other GI pathologies. The project recruited more than 400 pwCF, of whom more than 250 pwCF were assessed before and during a new treatment with ETI with the Patient Assessment of Constipation-Symptom severity Index (PAC-SYM) (29), the Patient Assessment of upper Gastrointestinal disorders-Symptom severity Index (PAGI-SYM) (30), and the Patient Assessment of Constipation-Quality of Life (PAC-QOL), as well as a modified Bristol Stool Scale (31). Whereas the authors found a significant ETI-induced decline in intestinal inflammation assessed by fecal calprotectin, regarding AS they report “statistically significant, but clinically unimportant changes in gastrointestinal symptoms in the PROMISE population” (27). Adjusted changes in PAC-SYM, PAGI-SYM and PAC-QOL scores only reached levels between 0.03 and 0.25 points, far below the minimal clinically important difference threshold, which, for PAC-SYM, has been considered to be about −0.60 (32).

These results are in stark contrast to three published studies using the CFAbd-Score^©^ and the CFAbd-day2day^©^ (its CF-GI-patient diary version). In patients older than 12 years, we recently found a significant and marked reduction in AS after ETI initiation (20, 33). Three different cohorts were assessed. Initially, a cohort of 107 pwCF from Germany and the UK followed up for 24 weeks, revealed a statistically significant and highly clinically relevant decline of 29% in total CFAbd-Scores^©^. At the same time, sub-scores for its five domains, namely “pain”, “GERD”, “disorders of bowel movement”, “disorders of appetite” and “GI-related QOL” decreased by 37%, 48%, 23%, 67% and 61%, respectively (all p<0.05) (8). Recently, these results were confirmed and shown to be sustained over 1, 2, 6 and 12 months on therapy with ETI in 108 pwCF included in the Irish/British RECOVER project. Declines in total CFAbd-Scores^©^ and its five domains showed significant decreases in a highly clinically relevant range during the mentioned period of time (10). Finally, these results were again confirmed using the diary version of the PROM, the CFAbd-day2day^©^, cumulatively calculating CFAbd-Scores^©^ after 2-4 weeks of therapy with ETI (20).

We attribute differences between the findings of both projects to the highly specific pattern of CF-related abdominal organ manifestations, including the frequent exocrine (and endocrine) pancreatic insufficiency, CFTR-related bowel pathologies, antibiotic-related dysbiosis (small bowel overgrowth SIBO), liver and bile pathologies. Furthermore, CF-specific effects of medications like pancreatic enzymes, nutrient supplementation, ursodeoxycholic acid, laxatives, as well as effects of varying adherence to the intake of these medications apparently lead to a very specific pattern of abdominal symptoms in pwCF (9). Consequently, even though non-CF-specific GI PROMs reveal an increase in the burden of abdominal symptoms in pwCF (22, 34), these questionnaires may not accurately capture the symptom pattern deriving from the mentioned CF specifics.

Accordingly, results obtained with the new CFAbd-Score.kid^©^ show significant decreases in AS during ETI therapy. We estimate that the improvement of 6.5 points (38%) in total CFAbd-Score.kid^©^ is likely to fall within the clinical relevant range, owing to the similarities observed in the changes achieved with the total CFAbd-Score^©^ in previous studies assessing ETI effects on AS, for which an estimate of the calculated minimal clinically important difference resulted in 3-4 out of 100 points (35). An accurate estimation of the minimal clinically important difference for the CFAbd-Score.kid^©^ is an issue of highest relevance, for which we are still collecting more data of cwCF from other CF centers, considering different age groups and other conditions that may provide more information about the burden of AS in pediatric cohorts. Additionally to the mentioned improvement in the total CFAbd-Score.kid^©^ significant improvements were found in mean sub-scores for the domains of “pain” (−37%), “QoL impairment” (−45%), “DBM/lower GI” (−31%) and “disorders of appetite” (−57%) (Fig. 2 and Tab. 2).

However, in contrast to our previous studies in pwCF aged ≥12 years, in children aged 6-11 years, we did not find a significant decline in the sub-scores for the GERD/upper GI domain during treatment. This may be due to the very low prevalence of GERD in children in the study at baseline, which agrees well to our very early findings during the development of the CFAbd-Score^©^ (19). Nonetheless, a tendency towards improvement is observed.

Interestingly, after ETI initiation, the number of patients in the present study reporting irregular “stool consistency” decreased by 24%. This contrasts with our previously published results obtained with the CFAbd-Score^©^ in pwCF aged ≥12 years, where improvement of stool consistency only reached 8% (8, 10, 20). This proportion, however, may be confounded with the mild adverse events of diarrhea, for which reported exposure-adjusted rates are slightly higher in pwCF aged ≥12 years (12.9% of patients; 31.94 events per 100 patient-years), compared to cwCF aged 6-11 years (10.6% of patients; 23.16 events per 100 patient-years) (36, 37).

In contrast to the percentage of pwCF ≥12 years of age reporting “fatigue”, little change was observed in the respective percentage of pediatric patients using the CFAbd-Score.kid^©^. Nevertheless, most of the included children reported a reduction in AS after ETI initiation. Only four of the 13 patients initially scoring below the first quartile Q_1_ of the CFAbd-Score.kid**^©^** baseline distribution, representing the lowest burden of symptoms, increased after ETI initiation. In two of them the total CFAbd-Score.kid^©^ result between Q_1_ and Q_2_, and two others even reached scores above Q_3_, after six months of ETI therapy. Indeed, it is expected that a lower percentage of patients may not benefit as much, or even display unexpected adverse effects induced by the highly effective therapy, similar to those reported regarding pulmonary function, sweat tests and weight gain (6). Enrolling more patients in further studies to assess symptom worsening would therefore be interesting.

Limitations of our study may include the irregularly separated time points at which the questionnaires were collected, partly owing to the SARS-CoV-2 pandemic. This prevented more detailed analyses as to when ETI effects on GI reach their maximum, or whether they plateau after some time. Furthermore, medication information such as laxatives and antibiotics was not consistently collected in all the participating CF Centers. These, as well as other open questions, will be the subject of future studies including a larger cohort and further relevant variables. Finally, item-by-item statistical comparisons between cwCF aged 6-11 years and pwCF ≥12 years of age were not conducted, as each age group only completed either the CFAbd-Score or the CFAbd-Score.kid, preventing a rigorous on-an-equal-footing comparison.

## 4 Conclusion

Using the novel CFAbd-Score.kid^©^ before and after ETI initiation in pediatric patients, we showed for the first time a significant reduction in the burden of abdominal symptoms in cwCF aged 6 through 11 years. This result demonstrates the newly developed PROM’s high “sensitivity to change”, e.g. to a therapeutic intervention, being a step in the validation process, following FDA guidelines. The CFAbd-Score.kid^©^ score has proved to be a useful tool for the assessment of AS in pediatric patients and will be further implemented in clinical practice and international studies. Given the cross-cultural issues, translating the CFAbd-Score.kid^©^ into other languages and extending the study to other European countries opens up avenues for further research (15).

## Data Availability

All data produced in the present study are available upon reasonable request to the authors

## Acknowledgements

we thank Daniel Kemp and Berit Mühl (CF-Selbsthilfe Dessau, Germany) for their substantial support and expertise in the development process of the CFAbd-Score.kid^©^. Furthermore, we thank Annette Kitzinger, developer and owner of Metacom^©^ (https://www.metacom-symbole.de/metacom_en.html) for providing the pictograms and symbols implemented in the pediatric version of the CFAbd-Score^©^. Finally, we would like to acknowledge the dedication and commitment of the children with CF and their parents who participated in this study, as well as the CF care teams at the study sites.

## Author contributions

Conceptualization: JGM, PS, LB, FD, CZ; Data curation CZ, PS, LB, LP; Formal analysis CZ, JGM, PS; Funding acquisition JGM; Investigation PS, LB, LP. LN, OE, SvD, UG-M; Methodology JGM, FD, CZ; Project administration JGM, FD; Resources JGM, LN, OE, UG-M; Software CZ; Supervision JGM, LN, OE, UG-M; Validation CZ, PS; Visualization; CZ, JGM, PS Writing - original draft CZ, JGM, PS; and Writing - review & editing JGM, CZ, PS, LB, FD, AB, LP, LN, OE, SvD, UG-M.

